# Expanding the spectrum of white matter abnormalities in Wolfram syndrome: A retrospective review

**DOI:** 10.1101/2024.08.31.24312796

**Authors:** Justin Simo, Heather M. Lugar, Elka Miller, Adi Wilf-Yarkoni, Yael Goldberg, Ayça Kocaağa, Shoichi Ito, Sirio Cocozza, Giulio Frontino, Cristina Baldoli, Aziz Benbachir, Catherine Ashton, Guy Rouleau, Tamara Hershey, Yann Nadjar, Roberta La Piana

**Affiliations:** The Neuro (Montreal Neurological Institute-Hospital), McGill University, Montreal, QC, Canada; Department of Neurology and Neurosurgery, McGill University, Montreal, QC, Canada; Department of Psychiatry, Washington University School of Medicine, St. Louis, MO, USA; Department of Diagnostic Imaging, The Hospital for Sick Children (SickKids), University of Toronto, Toronto, ON, Canada; Neuroimmunology Unit, Rabin Medical Center—Beilinson Hospital, Petach Tikva, Israel; Faculty of Medicine, Tel Aviv University, Tel Aviv, Israel; The Raphael Recanati Genetic Institute, Rabin Medical Center—Beilinson Hospital, Petach Tikva, Israel; Department of Medical Genetics, Eskisehir City Hospital, 71 Evler Mahallesi, Çavdarlar Sk., 26080, Odunpazarı/Eskişehir, Turkey; Department of Medical Education, Graduate School of Medicine, Chiba University, Chiba, Japan; Department of Advanced Biomedical Sciences, University of Naples “Federico II,” Naples, Italy; Department of Pediatrics, Diabetes Research Institute - IRCCS San Raffaele Hospital, Milan, Italy; Neuroradiology Unit, IRCCS San Raffaele Scientific Institute, Milan, Italy; Department of Neurology, Royal Perth Hospital, Perth, WA, Australia; Department of Human Genetics, McGill University, Montreal, QC, Canada; Mallinckrodt Institute of Radiology, Washington University School of Medicine, St. Louis, MO, USA; Department of Neurology, Pitié-Salpêtrière University Hospital, Paris, France; Department of Diagnostic Radiology, McGill University, Montreal, QC, Canada

**Author notes:** Correspondence: Roberta La Piana.

## Abstract

**Background and Objectives:** Wolfram syndrome (WFS) is a genetic disorder mainly caused by pathogenic variants in the *WFS1* gene. It is characterized clinically by optic atrophy (OA), diabetes mellitus (DM), sensorineural hearing loss (SNHL), diabetes insipidus (DI), and variable neurological/psychiatric symptoms. WFS typically manifests before age 20 and progresses into adulthood. Classical neuroradiological features include cerebellar and/or brainstem atrophy as well as white matter abnormalities ranging from small, ovoid lesions to diffuse, symmetrical changes along the visual pathway. Following the identification of multifocal, progressive white matter abnormalities that prompted the consideration of multiple sclerosis (MS) in two molecularly confirmed WFS subjects, we sought to verify whether MS-like lesions constitute a novel WFS-associated MRI pattern.

**Methods:** We conducted an international multicenter retrospective study of the clinical, genetic, and radiological data from 17 unrelated WFS subjects.

**Results:** Seven subjects (7/17; 41%) showed at least one focal white matter lesion evocative of MS. Among these seven, three fulfilled the McDonald radiological criteria of dissemination in space and time, suggesting an inflammatory demyelinating process. All subjects reviewed in the study had at least one of the classical WFS MRI features.

**Conclusions:** Our report expands the WFS spectrum of white matter involvement to include progressive, seemingly inflammatory demyelinating lesions. While we cannot exclude the possibility of a WFS-MS dual diagnosis in some cases, the role of *WFS1* in myelination suggests a selective white matter vulnerability in WFS. Our findings suggest that follow up MRI should be recommended to adult subjects with WFS. Further identification and longitudinal study of adult WFS subjects is required to confirm whether a WFS molecular diagnosis confers susceptibility to the development of MS.

## 1 Introduction

Wolfram syndrome (WFS) is a rare genetic spectrum disorder characterized clinically as any combination of optic atrophy (OA), diabetes mellitus (DM), diabetes insipidus (DI), and sensorineural hearing loss (SNHL), accompanied by other neurological and/or psychiatric features^1–4^. These latter features, seen in over 50% of cases, are usually progressive and include cerebellar ataxia, brainstem dysfunction, epilepsy, and cognitive disability^3^. Biallelic or monoallelic pathogenic variants in *WFS1* cause WFS, putatively through loss-of-function mechanisms that undermine endoplasmic reticulum (ER)-mitochondria dynamics and trigger unfolded protein response (UPR)-mediated cell death^5–11^.

From a neuroimaging perspective, although different CNS structures are involved, the most common imaging findings reported in WFS are cerebellar atrophy, brainstem atrophy, and signal changes along the visual pathway, posterior pituitary gland, pontine fibers, and cerebral white matter^12–15^. In particular, the patterns of white matter abnormalities range from small multifocal, ovoid-shaped T2-hyperintense lesions to symmetrical, diffuse involvement^12–17^. MRI findings in WFS can appear as early as the first decade, even when neurological symptoms are absent or barely noticeable, and they progress over time^3,9,10,12,18^. Interestingly, they seem to be coupled to more profound abnormalities that can be detected through advanced imaging techniques. These include microstructural alterations of the white matter in early myelinating structures (i.e., brainstem, posterior periventricular white matter), fueling the hypothesis that hypomyelination may play a role and precede adult stage neurodegeneration in WFS^9,10,14^.

In agreement with conventional and advanced neuroimaging findings that indicate white matter involvement in WFS, molecular studies strongly suggest that WFS1 could play a vital role in establishing and maintaining white matter integrity^17,19–24^. To this end, it has been speculated that the pathophysiology of CNS features in WFS could at least in part overlap that of more common demyelinating diseases, such as multiple sclerosis (MS)^14^. In line with these speculations, and after identifying two unrelated, genetically confirmed WFS subjects through the White Matter Rounds Network^25^, in which multifocal, progressive, and contrast-enhancing white matter abnormalities led to the consideration of MS, we aimed to assess the prevalence of likely inflammatory white matter lesions and MS diagnosis in subjects with WFS.

## 2 Materials and methods

### 2.1 Participants

We conducted an international, multicenter retrospective review of the clinical, neuroradiological, and molecular data of subjects with the following inclusion criteria:

1. Clinical diagnosis of WFS, defined as the presence of:

a. At least <u>two</u> of the four classical WFS features (OA, DM, DI, and SNHL).
b. At least <u>one</u> neurological or psychiatric symptom in addition to SNHL and OA. Patients with Wolfram-like syndrome (at least <u>one</u> classical feature plus one neurological/psychiatric symptom aside from SNHL and OA) were included if they had a positive WFS family history.
2. If above criterion not met, genetic diagnosis molecularly confirmed by detection of pathogenic or likely pathogenic variants in the *WFS1* gene, classified according to ACMG guidelines^26^, through an accredited clinical diagnostic laboratory.
3. Available brain MRI with axial and sagittal T2-weighted and T2-FLAIR images at minimum. Any additional brain MR sequences or spinal cord images for subjects meeting this minimum requirement were also reviewed.

To achieve this, we contacted the corresponding authors of previously published WFS case reports and series (150 subjects). Ten authors responded and agreed to participate. We therefore included 17 subjects from seven countries: Canada (1), France (1), Israel (2), USA (8), Italy (3), Turkey (1), and Japan (1).

### 2.2. Data Analysis

For each included subject, we collected the following demographic and clinical data: sex, family history, age at onset (defined as the age at which at least one WFS classical feature or neurological/psychiatric deficit appeared), age at latest examination, and presence of ataxia/cerebellar signs, spasticity, abnormal reflexes, other movement disorders, cranial nerve dysfunction, vertigo, tinnitus, migraine, neuropathy, seizures, psychiatric disorders, cognitive deficits, sphincter dysfunction, and extra-neurological signs.

The MRI images were reviewed independently by two teams of raters:

a. JS and RLP, with >15 years of experience in white matter disorders.
b. A neuroradiologist from each participating institution (SC/GF-CB for cases from Italy, AWY-YG for cases from Israel, AK for the case from Turkey, and SI for the case from Japan) **<u>or</u>** a neuroradiologist from outside the study (EM, with >20 years of experience, for all cases from Canada, USA, and France).

When available, for 10 subjects (10/17; 58.8% of the entire cohort), each MRI scan was re-evaluated four months later, blind to the initial MRI interpretation, with a focus on the multifocal white matter abnormalities documented in each subject. The raters remained blind to the clinical data during each review session to further mitigate lesion classification bias. Disagreements between raters were resolved by consensus.

We performed a qualitative analysis of all available MRI images to assess the presence and characteristics of the white matter abnormalities. The following criteria were analyzed: location (periventricular, subcortical, juxtacortical), lobar involvement (frontal, temporal, parietal, occipital, cerebellar), pattern of involvement (multifocal, confluent, diffuse), symmetry, and enhancement following gadolinium injection. In our study, a lesion was defined as a focal area of T2-hyperintense signal in the CNS white matter. Juxtacortical lesions were defined as lesions in direct contact with the cortical ribbon. Periventricular lesions included all lesions abutting any portion of lateral ventricles as well as intracallosal lesions and linear plate-like bandings. Subcortical lesions were defined as supratentorial non-juxtacortical, non-periventricular white matter lesions. MS-like lesions were defined as round/ovoid areas of T2-hyperintense signal at least 3mm in long axis^27–29^. MR images were also evaluated for cerebral, brainstem, and cerebellar (hemispheric and vermian) atrophy and when possible, depending on the available sequences, other associated features such as gray matter involvement, microvascular changes, and calcifications. When spinal cord MRI was available, we evaluated it for the presence of signal abnormalities and atrophy. Given the nature of this retrospective review, there were intrinsic limitations in the analysis, as MRI examinations slightly differed in imaging quality and techniques.

### 2.3 Standard Protocol Approvals, Registrations, and Patient Consent

Access to patient medical histories and results of diagnostic investigations for collaborative retrospective study was granted by the review boards of each participating institution, in conformity to respective anonymization procedures.

## 3 Results

**Table 1** contains the clinical, genetic, and radiological data from the 17 included subjects.

**Table 1.** Clinical, genetic, and neuroradiological data compiled from 17 WFS subjects. Legend: A = ataxia and/or other cerebellar signs; B = bladder dysfunction; BA = brainstem atrophy; C = cataract; CA = cerebellar atrophy; Cal = callosal/pericallosal; CD = cognitive deficit; Cer = cerebellar white matter/peduncles; C Het = compound heterozygous; D = diffuse/confluent involvement of the centrum semiovale and/or peritrigonal white matter; DCo = diabetic coma; DI = diabetes insipidus; DM = diabetes mellitus; DN = diabetic neuropathy; Het = heterozygous; Hom = homozygous; Hyperr = hyperreflexia; Hypor = hyporeflexia; JxU = juxtacortical/U-fiber; LP = lumbar puncture; M = migraine; Mo = other motor issues; MS = multiple sclerosis; N/A= not available; N/P = not performed; OA = optic atrophy; OCBs = oligoclonal bands; Pit = absent/diminished posterior pituitary bright spot; PN = peripheral neuropathy; Po = pontine signal changes; Psy = psychiatric symptoms; Pun = punctate subcortical white matter foci typical of WFS; PV = periventricular; PVp = diffuse, bilateral signal changes in posterior horn periventricular white matter and/or optic radiations; S = seizures; SC = spinal cord; SNHL = sensorineural hearing loss; Sp = spasticity; Sub = subcortical; WBCs = white blood cells; † = focal lesion(s) ultimately categorized as intermediate (in between WFS and MS); ** = McDonald radiological criteria fulfilled.

| Subject<sup>ref</sup> | Origin | Age at onset/last exam (decade) | Zygosity | WFS1 variants (NM_006005.3) | Classic WFS symptoms | Neurological symptoms | Age at last MRI (decade) | Classical WFS MRI features | MS-like white matter foci | Dissemination in space and/or time | Gadolinium enhancement | Additional investigations or interventions | Full McDonald criteria (clinical + radiological) |
| --- | --- | --- | --- | --- | --- | --- | --- | --- | --- | --- | --- | --- | --- |
| 1 | Canada | 30s/40s | Het | c.2389G>A, p.D797N | SNHL, OA | Hypor, M, PN | 40s | Pit, PVp, Pun | Sub, PV/Cal, JxU | Yes, space + time** | N/P | N/P | Partially fulfilled |
| 2 | France | 10s/30s | C Het | c.1112G>A, p.W371*<br>c.1511C>G, p.P504R | DM, Vision loss | Hyperr, A, B | 20s | D | Sub, PV/Cal, SC | Yes, space + time** | Yes (Cal; Sub) | LP: inflammatory profile w/ OCBs | Fulfilled |
| 3 | Israel | 10s/30s | Hom | c.1230_1233delCTCT, p.V412Sfs | SNHL, OA, C | A, Hypor, S | 30s | CA, PVp, D | Sub, JxU, PV, Cer, SC | Yes, space + time** | Yes (Sub – parietal) | LP: elevated WBCs w/ OCBs; MS-like foci stabilized upon teriflunomide use, while non-focal abnormalities progressed | Fulfilled |
| 4<sup>18</sup> | Israel | 10s/60s | Hom | c.1672C>T, p.R558C | DM, OA | A, CD, B | 60s | D, PVp | Sub | Yes, time only | Yes (Sub – occipital) | N/P | Partially fulfilled |
| 5<sup>45</sup> | Italy | Unknown/50s | N/A | N/A | N/A | N/A | 50s | CA, BA, PVp | - | - | N/P | N/P | - |
| 6<sup>46</sup> | Japan | 0s/30s | N/P | N/P | DM, SNHL, OA | DCo | 30s | CA, BA, Pit, D, PVp | - | - | N/P | N/P | - |
| 7<sup>14</sup> | USA | 10s/20s | C Het | c.817G>T, p.E273*<br>c.1839G>A, p.W613* | DM, DI, SNHL, OA | A, Mo, S, Psy, CD, B | 10s | Po, PVp, Pun† | PV | - | N/P | N/P | - |
| 8<sup>14</sup> | USA | 10s/20s | C Het | c.605A>G, p.E202G<br>c.631G>A, p.D211N | DM, OA | A | 20s | CA, PVp (unilateral), Pun | Sub† | - | N/P | N/P | - |
| 9<sup>14</sup> | USA | 10s/20s | C Het | c.376G>A, p.A126T<br>c.1838G>A, p.W613* | DM, DI, OA | A, Psy, B | 20s | Po, PVp, Pun | - | - | N/P | N/P | - |
| 10<sup>14</sup> | USA | 20s/30s | C Het | c.1240_1242del, p.P414del<br>c.1698_1703del, p.L567_P568del | DM, DI, SNHL, OA | A, Mo, Psy, B | 30s | CA, BA, PVp, Pun | PV | - | N/P | N/P | - |
| 11<sup>14</sup> | USA | 20s/20s | C Het | c.320G>A, p.G107E<br>c.1885C>T, p.R629W | DM, DI, SNHL, OA | A, Mo, S, B | 20s | CA, BA, Po, PVp, Pun | - | - | N/P | N/P | - |
| 12<sup>14</sup> | USA | 10s/10s | C Het | c.740_741del, p.P247Cfs<br>c.1243_1245del, p.V415del | DM, DI, SNHL, OA | A, Mo, M, PN, Psy, B | 10s | CA, PVp, Pun | - | - | N/P | N/P | - |
| 13<sup>14</sup> | USA | 10s/20s | C Het | c.599T>C, p.L200P<br>c.695G>C, p.R232P | DM, SNHL, OA | A, Sp, Psy, CD, B | 20s | BA, Po, PVp, Pun† | Sub, PV | - | N/P | N/P | - |
| 14<sup>14</sup> | USA | 10s/10s | C Het | c.1251_1252delinsG, p.P417Lfs<br>c.1885C>T, p.R629W | DM, DI, SNHL, OA | Hypor, Psy, B | 10s | PVp, Pun | - | - | N/P | N/P | - |
| 15<sup>17</sup> | Turkey | 10s/10s | Het | c.1230_1233delCTCT, p.V412Sfs | OA, C | A, S, CD | 10s | BA, Po, D | - | - | N/P | N/P | - |
| 16 | Italy | 0s/20s | C Het | c.605A>G, p.E202G<br>c.1628T>G, p.L543R | DM, OA | DN, B | 30s | Po, PVp | - | - | N/P | N/P | - |
| 17 | Italy | 0s/10s | C Het | c.409_424dup16, p.V142Gfs<br>c.1628T>G, p.L543R | DM, DI, SNHL, OA | Psy, B | 10s | Po, PVp | - | - | N/P | N/P | - |

### 3.1 Clinical findings

Demographic and clinical data were available for 16/17 subjects (94.1%); for one subject the only information was the sex, the age at last examination, and the confirmed molecular diagnosis. The cohort included nine female subjects (52.9%). Median age at onset of WFS was 15.5 years (range = 3-35 years). Regarding classical WFS features, nearly all exhibited OA (n = 15/16; 94%) except for one subject (#2), who had diminished visual acuity with a normal fundoscopic examination. Thirteen had DM (n = 13/16; 81%), nine had SNHL (n = 9/16; 57%), and seven had DI (n = 7/16; 44%). In all subjects except subject #11, for whom the first sign was non-syndromic epilepsy, the first sign(s) included one or multiple classical WFS features (n = 15/16; 94%), most commonly DM. Two subjects, #3 and #15, developed cataract, an uncommon, yet known WFS feature, at some point in the clinical course (n = 2/16; 13%).

Neurological and/or psychiatric symptoms, aside from OA and SNHL, manifesting several years post-onset (median = 6 years; range = 3-42 years), were documented in all subjects (n = 16/16; 100%), consistent with the neurodegenerative character of WFS.

Two subjects, #2 and #3 (n = 2/16; 13%), presented acute focal deficits in the third/fourth decade that may be related to active demyelination as shown on MRI. These included hyperreflexia and fine motor impairment (subject #2) and truncal ataxia, appendicular ataxia, and dysarthria (subject #3).

### 3.2. Genetic findings

Pathogenic or likely pathogenic variants in *WFS1* (mono-or biallelic) were documented in all subjects for whom genetic testing had been performed (n = 16 tested/17 total; 94%). Subject #15 was diagnosed with Wolfram-like syndrome after detection of a monoallelic pathogenic *WFS1* variant through whole genome sequencing, which was performed because of unusual MRI features.

Family history was noteworthy for symptoms along the WFS spectrum in most subjects for whom this information was available (n = 10/15; 66%). The family history was positive for WFS in three cases (n = 3/15; 20%), cataract in three (n = 3/15; 20%), and cardiac anomalies/syndromes in two (n = 2/15; 13%), which have also been reported in WFS.

### 3.3. Neuroimaging findings

The mean age at the latest MRI available for review was 29 years old (range = 14-60 years). All subjects demonstrated at least one neuroradiological feature typical of WFS (**Figure 1**)^12–15^. For 11 subjects (n = 11/17; 65%), two timepoints were available for assessment (mean follow-up MRI duration = 5.3 years).

**Figure 1.**
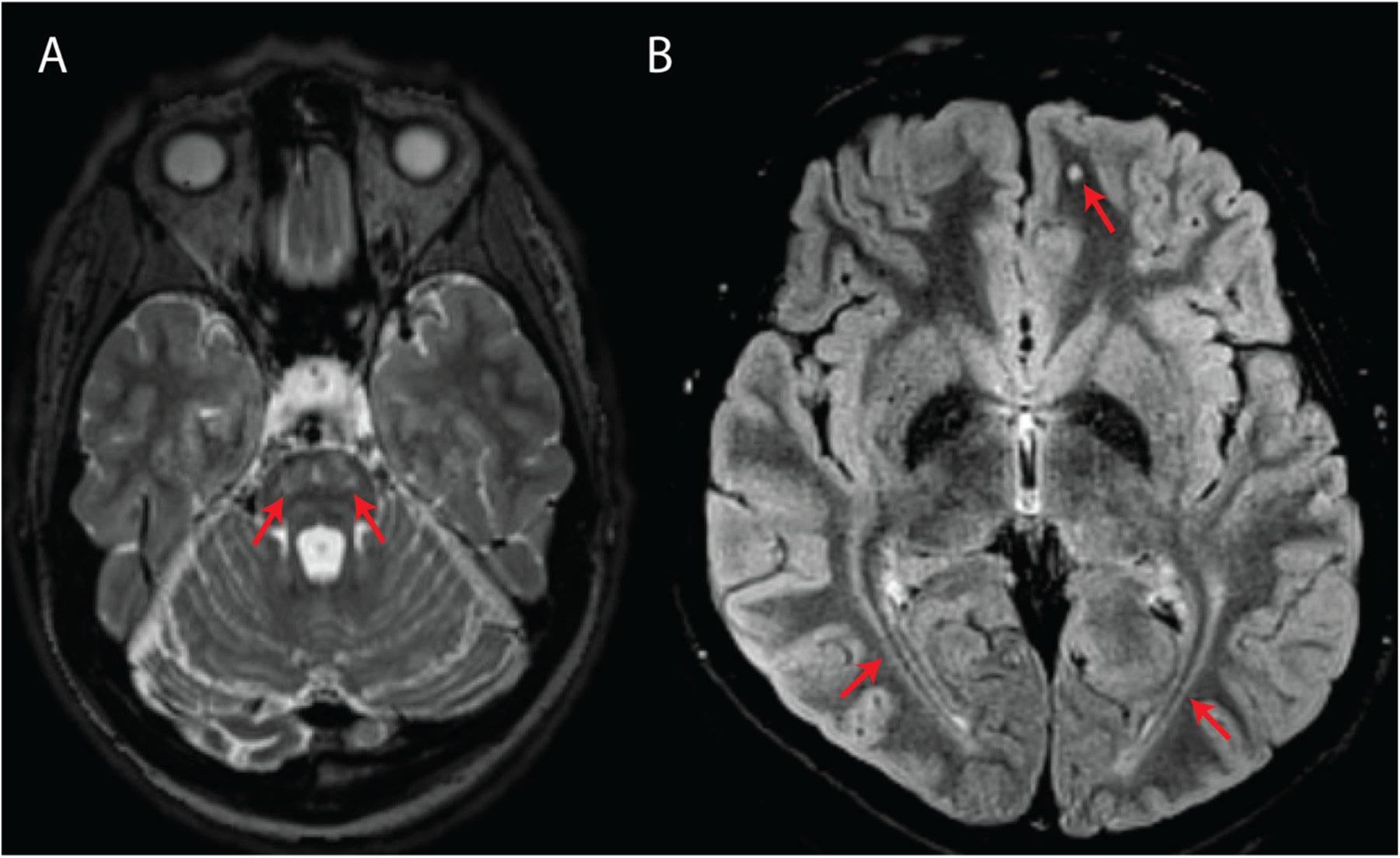
Classical MRI features associated with WFS. **A**, Axial T2-weighted image showing bilateral pontine signal abnormality (subject #10). **B**, Axial T2-FLAIR image showing bilateral symmetric posterior white matter abnormal signal as well as a punctate, round white matter signal abnormality in the left frontal lobe (subject #14).

#### Classical WFS-associated features

Involvement of the posterior periventricular white matter and/or optic radiations was the most common finding (n = 15/17; 88%), followed by punctate subcortical white matter changes (n = 9/17; 53%), pontine signal changes (n = 7/17; 41%), brainstem atrophy (n = 6/17; 35%), cerebellar atrophy (n = 5/17; 29%), diffuse/confluent involvement of the centrum semiovale and/or peritrigonal white matter (n = 4/17; 24%), and absent/diminished T1-weighted posterior pituitary bright spot (n = 1/17; 6%).

Enlarged perivascular spaces (n = 3/17; 18%), corpus callosum abnormalities (n = 3/17; 18%), midbrain signal changes (n = 2/17; 12%), and abnormal occipital lobe gyrification with or without posterior horn ventricular enlargement (n = 2/17; 12%) were additionally observed.

#### MS-like white matter abnormalities

We detected T2-hyperintense white matter foci that were MS plaque-like^27–29^ in seven subjects (n = 7/17; 41%) (**Figures 2 and 3**). In four of these subjects (n = 4/17; 24%), the McDonald radiological criteria of dissemination in space and time were either fully (n = 3) or partially (n = 1) fulfilled, inciting consideration of a WFS-MS double diagnosis (**Figure 2**)^27^. Some lesions in three of these four subjects showed gadolinium enhancement on T1-weighted imaging (**Figure 2A**), further supporting the existence of an underlying inflammatory process. Positive oligoclonal band status strongly supported a secondary diagnosis of MS in the only two subjects for whom lumbar puncture was performed (#2 and #3). In both cases, a secondary diagnosis of MS was communicated, and MS treatment with teriflunomide was initiated in subject #3. Follow-up MRI after teriflunomide initiation documented stabilization of the multifocal white matter lesion load with worsening of pre-existing cerebellar atrophy and diffuse bilateral subcortical white matter involvement (**Figure 2G-I**). The observed locations and characteristics of MS-like focal lesions are summarized in **Table 2**.

**Figure 2.**
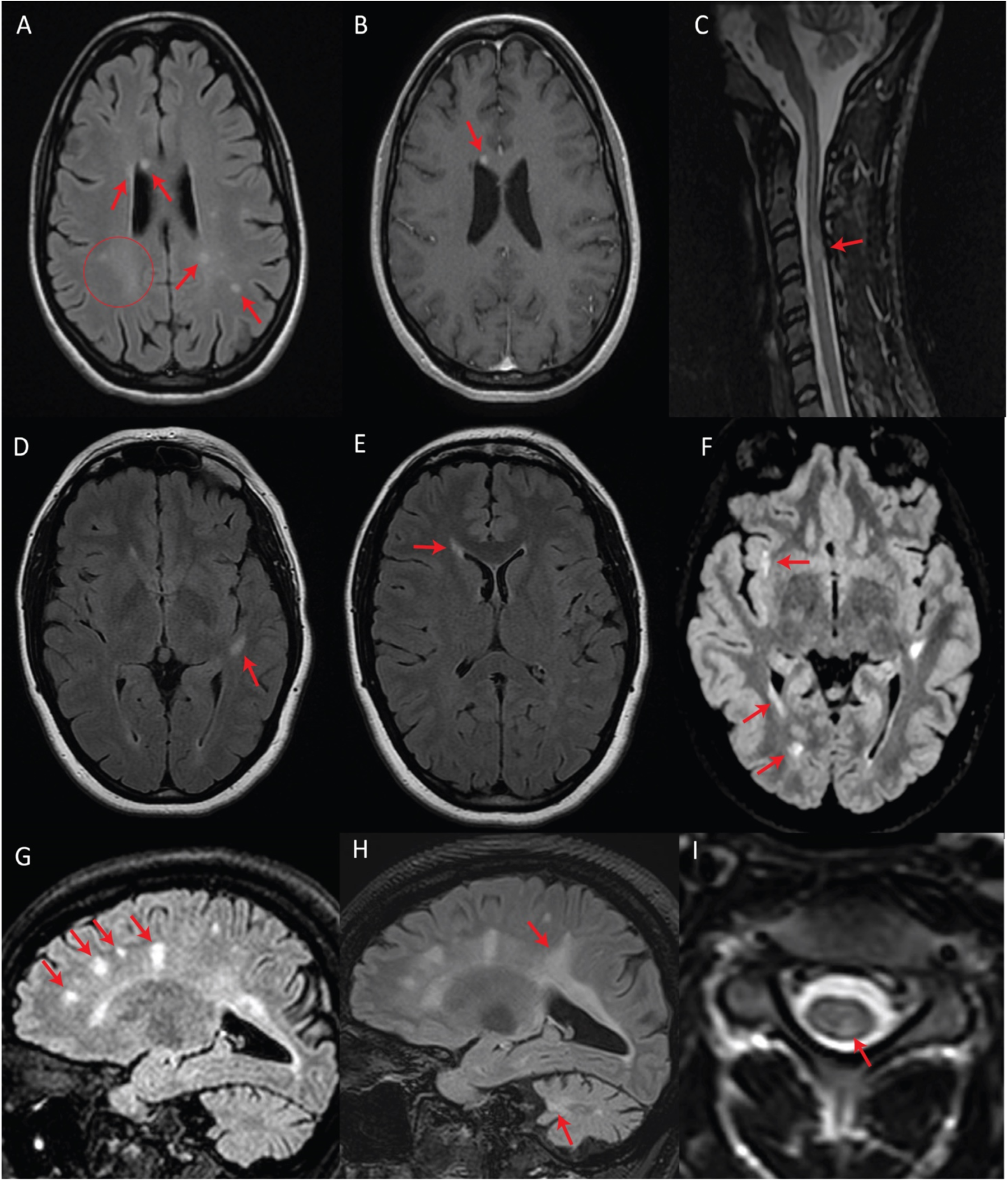
Patients with fulfilled or partially fulfilled McDonald criteria. **A**, **B**, and **C** represent subject #2 (criteria fulfilled). **A**, Axial T2-FLAIR image showing multifocal periventricular, callosal, and subcortical white matter lesions (arrows) with accompanying diffuse abnormal peritrigonal white matter signal (circle). **B**, Post-gadolinium axial T1 image showing an enhancing callosal lesion. **C**, Sagittal T2 image of the cervical and thoracic spinal cord showing a demyelinating lesion at the level of C3. **D**, **E**, and **F** represent subject #1 (criteria partially fulfilled). **D**, Axial T2-FLAIR showing an oval-shaped lesion in the left temporal subcortical white matter. **E**, Axial T2-FLAIR showing a periventricular lesion around the frontal horn of the right lateral ventricle. **F**, Axial T2-FLAIR, performed seven years after the first exam (**D**, **E**), showing new focal white matter lesions in the right insula, right periventricular white matter, and along the posterior horn of the right lateral ventricle. **G**, **H**, and **I** represent subject #3 (criteria fulfilled). **G**, Sagittal T2-FLAIR showing multifocal white matter lesions perpendicular to the ventricular walls. **H**, Sagittal T2-FLAIR, performed four years after initial MRI (G), demonstrating progression of the lesion burden in both the supra- and infratentorial regions despite treatment with teriflunomide as of the first year in the four-year interval. **I**, Axial T2 spinal cord MRI showing a white matter lesion located in the left posterior cord at the C3 level.

**Figure 3.**
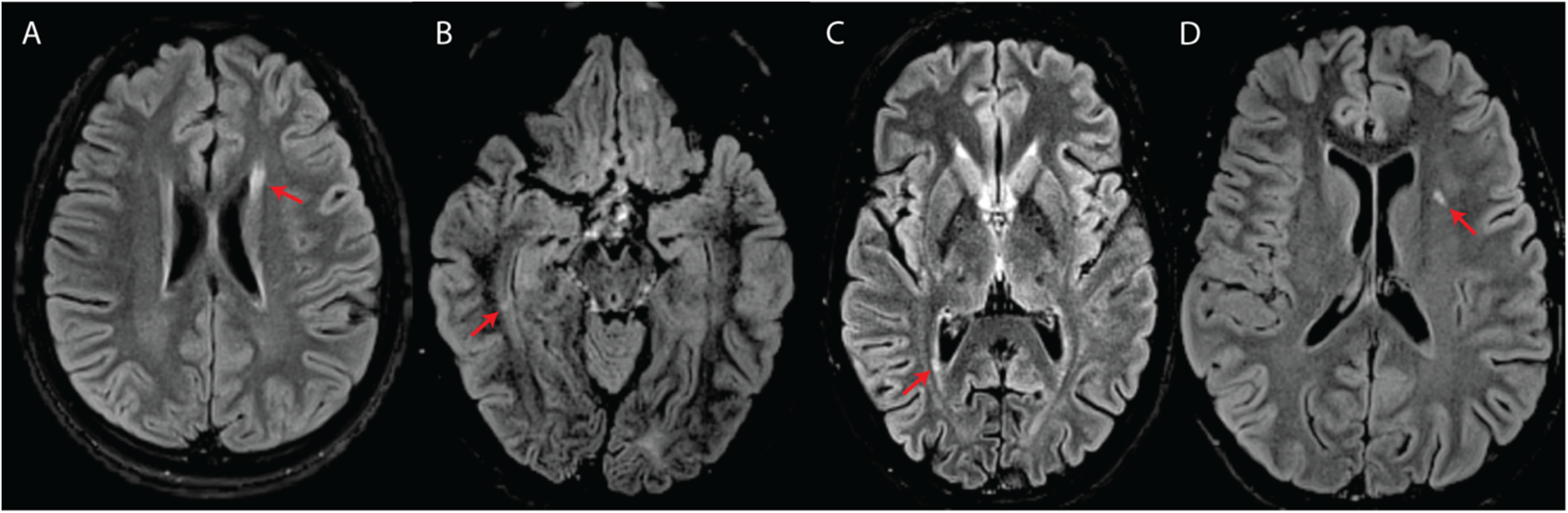
White matter abnormalities suggestive of an inflammatory process. (**A**, **B**, **C**) or classified as intermediate (**D**) in four subjects with WFS. All images are axial T2-FLAIR. **A**, Elongated lesion located at the level of the anterior horn of the left lateral ventricle (subject #7). **B**, Small temporal periventricular lesion perpendicular to the right ventricular wall (subject #7). **C**, Small periventricular lesion perpendicular to the right ventricular wall (subject #10). **D**, Left insular deep white matter lesion classified as intermediate between suspicious for inflammatory process and typical for WFS (subject #8).

**Table 2.**
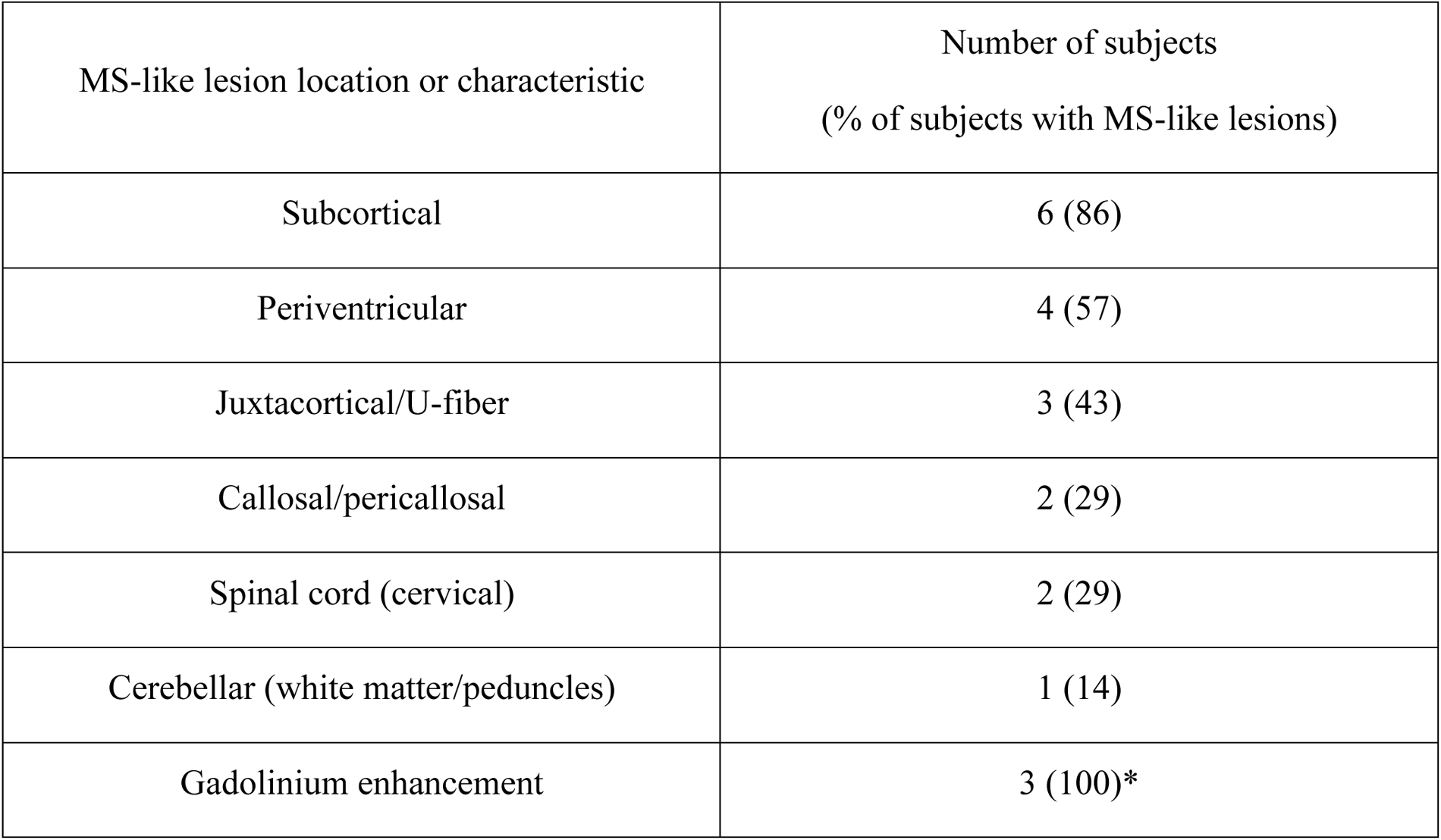
Distribution of MS plaque-like foci across the seven subjects in which these were identified. *Only three radiologically MS-like subjects underwent post-contrast imaging.

During blind re-analysis four months after the first image review, it was agreed that three subjects (#7, #8, and #13) had focal lesions not strictly belonging to either category (WFS- or MS-like/demyelinating) (**Figure 3D**; see **Table 1** for initial lesion categorization in each subject). The areas of suspicion were in the subcortical white matter and included two or more small, round, yet non-ovoid T2-hyperintense lesions close to one another. The lesions did not correspond to enlarged perivascular spaces.

## 4 Discussion

In this study, we report WFS subjects presenting with neuroradiological and, in some cases, clinical profiles compatible with a diagnosis of MS. The identification of four subjects with seemingly inflammatory white matter lesions that fully or partially fulfilled the McDonald criteria for MS diagnosis raises the possibility that a WFS molecular diagnosis may predispose to the development of neuroinflammatory white matter disorders such as MS. In fact, the worldwide prevalence estimates of MS (∼1:2800)^30^ and WFS (∼1:500 000)^31,32^ argue against coincidental occurrence of WFS and focal demyelinating events but rather suggest pathophysiological overlap between MS and WFS.

Our series focuses on MR imaging of adult subjects with WFS, who have been far less studied by imaging than pediatric subjects. Our findings suggest that secondary neuroinflammation, even when it does not satisfy the full criteria for MS, might contribute to gradual neurological and/or psychiatric decline in WFS^9^. Therefore, we recommend follow-up of WFS patients beyond adolescence to monitor the onset of inflammatory demyelinating lesions.

Importantly, multifocal white matter lesions have been previously described in WFS, albeit not with the usual features of MS plaques. Thus, the risk for individuals with Wolfram-like syndrome (i.e., partial WFS) to receive an incorrect primary diagnosis of MS is not negligible given the variability in neurological progression seen across subjects. We recommend that genetic testing for WFS be performed when a subject presents with multifocal white matter lesions – either seemingly inflammatory or not – and at least one of the classic diagnostic features of WFS.

In line with the hypothesis that some subjects in our study may be concomitantly suffering from WFS and MS, subject #3 responded to teriflunomide (**Figure 2G-I**). This was the only subject in our series who had started disease-modifying therapy for MS. While there is a possibility that the treatment had no impact on the natural course of WFS in this subject, it is plausible that underlying degenerative processes may have been exacerbated while inflammation was being suppressed. Even though the exact mechanisms contributing to this subject’s outcome remain undefined, simultaneous wolframin/WFS1 deficiency and teriflunomide treatment might have imposed a significant mitochondrial stressor. Indeed, Wolframin/WFS1 plays an indirect, yet essential role in multiple facets of mitochondrial quality control putatively through modulation of Ca^2+^ homeostasis, while the mechanism of action of teriflunomide is the inhibition of dihydroorotate dehydrogenase, whose activity is intrinsically linked to the mitochondrial respiratory chain^19,33^. Thus, the classic WFS-associated neuroradiological features in this subject might have progressed, at least in part, because of a compounded mitochondrial insult, although dedicated studies are needed to validate this speculation.

Existing molecular evidence insists that unchecked tissue-specific and systemic inflammatory processes are significant drivers of WFS pathogenesis^34–38^. For example, in the context of DM, several works suggest that loss of wolframin protein (Wfs1) in murine pancreatic β-cells causes ER stress-mediated NLRP3 inflammasome activation and subsequent apoptosis by means of upregulated expression of pro-inflammatory cytokines, namely IL-1β^35,37,38^. β-cells lacking wolframin in turn show disproportionate increases in ER stress and pro-inflammatory mediator expression in response to applied cytokines and hyperglycemia, forming a positive feedback cycle that likely exacerbates DM in WFS^35^. In the context of OA, early stage optic nerve demyelination in *Wfs1*-KO mice is marked by ER stress-mediated STAT3 activation and an overall pro-neuroinflammatory, anti-survival transcriptomic shift in local glial populations^34^. This shift is associated with a downregulation of glial MCT1 that hampers neuron-bound lactate shuttling capacity, instigating retinal ganglion cell degeneration over time^34^. Critically, lower expression of MCT1 was additionally detected in the cortex and striatum of the mice, indicating that these neuroinflammatory transcriptomic shifts may tile the entire brain^34^. Furthermore, in a recent WFS case report, patient-derived peripheral blood mononuclear cells (PBMCs), which were anomalously wolframin-deficient, showed constitutively high, UPR-independent expression of pro-inflammatory cytokines that correlated with hypercytokinemia (TNF-α, IL-1β, and IL-6) and a positively skewed T_h_17/T_reg_ ratio—signs of systemic inflammation^37^. A chronic, systemic inflammatory state characterized by hypercytokinemia was also corroborated in a WFS mouse model^35^. Such findings are particularly intriguing given the large body of evidence implicating T_h_17 cells in MS pathogenesis, importantly during the acute phase of disease^39^. Taken together, these studies implicate wolframin in the regulation of parallel inflammatory signaling cascades that are perhaps simultaneously brewing in a WFS individual. Our result of focal enhancement post-contrast being observed in all radiological MS-mimicking subjects for whom this investigation was performed (subjects #2, #3, and #4), when interpreted alongside a potential chronic T_h_17-driven systemic inflammatory process and generalized neuroinflammatory state, begs the question of whether MS-overlapping pathology could arise in WFS.

In our review, some lesions were interpreted as neither MS-nor WFS-like. In the corresponding subjects, gadolinium administration was not performed, so we do not know whether these lesions could be inflammatory. This observation further supports the idea of a continuum between white matter involvement classically seen in WFS and in neuroinflammation. The identification of signs currently considered specific for MS, namely the central vein sign (CVS), could help in further classifying white matter lesions in WFS^40^. CVS could not be validated in our subjects due to a lack of susceptibility-weighted imaging sequences, no reconstructible planes, and probably insufficient field strength. Higher field imaging (7T MRI) could prove useful in not only detecting CVS, as demonstrated by a recent study evaluating the diagnostic performance of 3T vs. 7T MRI in MS^41^, but also in better depicting the signal characteristics of white matter abnormalities in WFS subjects, which may help differentiate WFS from MS by imaging.

A recent prospective study has supported the addition of the optic nerve as a fifth CNS location within the McDonald radiological criterion of dissemination in space^42^. Since dedicated optic nerve MRI was only performed in one subject from our cohort (who did not have a confirmed molecular diagnosis), and as mentioned, adult WFS subjects have been far less studied than pediatric subjects, the prevalence of optic nerve lesions in the adult WFS population remains unknown. Consequently, the proportion of WFS patients who will satisfy MS diagnostic criteria because of the proposed modification to the criteria cannot be estimated at this time.

Our study has several limitations, namely its retrospective nature (which precluded the use of advanced imaging techniques), the obstacles related to the identification and inclusion of adult WFS patients, and the lack of follow-up MRI in a considerable number of subjects. Future prospective imaging studies can help clarify the nature of white matter involvement in adult patients with WFS.

In conclusion, our report expands the WFS spectrum of white matter involvement to include late-onset progressive, seemingly inflammatory demyelinating lesions that, in the context of supportive clinical and laboratory findings, can prompt consideration of MS. We emphasize the need for continuous radiological follow-up of WFS patients into early and middle adulthood, and the utility of international collaboration in this regard, for better characterization of age- and/or immune-related MRI patterns. In the future, these results may lead to the inclusion of WFS to the growing list of single-gene disorders with the potential to mimic or confer susceptibility to MS^43,44^.

## Data Availability

All data produced in the present study are available upon reasonable request to the authors.

## 6 Author Contributions

JS: manuscript writing and project coordination (contacting WFS authors, collection, compilation, and analysis of data, via conference call or email correspondence, from all authors who accepted to collaborate). EM: external neuroradiologist invited for MRI review. HML & TH: data collection from WFS cohort followed at Washington University, USA (eight subjects). AWY & YG: data collection from two WFS subjects followed at Tel Aviv University, Israel. AK: data collection from one Wolfram-like syndrome subject followed at Eskisehir City Hospital, Turkey. SI: data collection from one WFS subject followed at Chiba University, Japan. SC: data collection from one WFS subject seen at University of Naples “Federico II,” Italy. GF & CB: data collection from two subjects, derived from a larger cohort of 16 pre-screened WFS subjects (of which six were adults) seen at San Raffaele Hospital, Italy. AB: analysis of compiled data. CA, GR, & RLP: data collection from one WFS subject followed at McGill University, Canada. YN: data collection from one WFS subject followed at Pitié-Salpêtrière University Hospital, France. RLP: conceptualization of the study, analysis of compiled data, funding acquisition, and review and editing of the manuscript.

## 8 Acknowledgment

This work would not have been possible without our international collaborators, who are members of the [White Matter Rounds Network](https://www.whitematterrounds.com/en/):

*“Adult genetic leukoencephalopathies are rare neurological disorders that present unique diagnostic challenges due to their clinical and radiological overlap with more common white matter diseases, notably multiple sclerosis (MS). In this context, a strong collaborative multidisciplinary network is beneficial for shortening the diagnostic odyssey of these patients and preventing misdiagnosis. The White Matter Rounds (WM Rounds) are multidisciplinary international online meetings attended by more than 30 physicians and scientists from 15 participating sites that gather every month to discuss patients with atypical white matter disorders. […] The WM Rounds Network demonstrate[s] the value of collaborative multidisciplinary international case discussion meetings in differentiating and preventing misdiagnoses between genetic white matter diseases and atypical MS.”*

- Huang YT, et al. The White Matter Rounds experience: The importance of a multidisciplinary network to accelerate the diagnostic process for adult patients with rare white matter disorders. *Front. Neurol.* (2022) 13:928493. doi: 10.3389/fneur.2022.928493

